# Timeliness of Yellow Fever Specimen Collection and Transport in Ghana, 2018-2022

**DOI:** 10.1101/2025.06.19.25329877

**Authors:** Seth D. Judson, Lee Schroeder, Franklin Asiedu-Bekoe, Dennis Odai Laryea, Gifty Boateng, Horlali Gudjinu, Robert Ossom, Jerry Fosu Danquah, David W. Dowdy, Ernest Kenu

## Abstract

Yellow fever is a mosquito-borne viral hemorrhagic fever that has caused recent outbreaks in African countries, including Ghana (2021-2022). Delayed diagnosis of yellow fever may cause increased morbidity and mortality. To improve timely detection of yellow fever, we need to better understand the factors contributing to diagnostic delays. We analyzed the diagnostic testing timeline of all suspected yellow fever cases in Ghana from 2018-2022. For these patients we calculated the days from symptom onset to specimen collection and arrival at the National Public Health and Reference Laboratory (NPHRL) for testing. We compared these times to World Health Organization (WHO) metrics. For suspected yellow fever cases, the time from symptom onset to specimen arrival had a median of 10 days (interquartile range 6-17). 5892/6345 (93%) of specimens were collected within 14 days of symptom onset, and 2653/6471 (41%) of specimens arrived within 3 days of collection (WHO metrics). Overall, we find that the timing of yellow fever testing varies among districts in Ghana. While specimens are generally collected in a timely manner, delays in specimen arrival are common. Improving specimen transport for yellow fever and/or expanded testing could lead to more timely detection of outbreaks.

## Introduction

Multiple recent yellow fever (YF) outbreaks have occurred in Africa, and delays in diagnosing and implementing control measures for YF have been associated with increased morbidity and mortality [1,2]. While an effective vaccine for YF exists, there are ongoing gaps in immunization coverage [3]. Additionally, constraints on diagnostic resources and healthcare systems contribute to delayed detection of YF outbreaks [4].

Ghana is classified as a high-risk country for YF by the World Health Organization (WHO) and has had multiple yellow fever outbreaks, including a recent YF outbreak from 2021-2022 [5–7]. Recent yellow fever outbreaks in Ghana have originated in remote northern districts among unvaccinated nomadic populations that are far from the NPHRL that performs YF testing, potentially increasing the time to outbreak detection [5,8].

Yellow fever virus is transmitted among humans in rural and urban settings in Africa by *Aedes* mosquitoes [5,9]. It is estimated that roughly half of patients with YF are asymptomatic, while approximately 15% of patients develop severe disease with an estimated case fatality rate of 30-50% [10,11]. Given the overlap of YF symptoms with other infectious diseases such as malaria and viral hepatitis, clinical diagnosis of YF is challenging and thus laboratory diagnosis is essential for surveillance and outbreak detection [12,13]. Early diagnosis of YF is important for supportive care and outbreak containment measures including vector control, risk communication, patient isolation, and reactive vaccination [2,3].

When a patient meets the suspected case definition for YF in Africa, a blood specimen must be collected and sent to a national reference laboratory for initial testing, usually consisting of a YF IgM antibody capture enzyme-linked immunosorbent assay (MAC-ELISA) [14–16]. There is potential for false positive results with the YF IgM MAC-ELISA from cross-reactive antibodies from other co-circulating flaviviruses (such as dengue or Zika viruses) or from recent YF vaccination [12,13]. Confirmatory testing involves reverse transcriptase polymerase chain reaction (RT-PCR) or plaque reduction neutralization testing (PRNT), and specimens must be shipped to regional reference laboratories or specialized laboratories to perform these tests. If a blood specimen is collected within 14 days from symptom onset, YF RNA in the specimen may be detectable via RT-PCR (although the probability decreases after 10 days), otherwise PRNT has to be performed which is far more time-intensive and requires a BSL-3 facility [13,14].

There are multiple steps in the YF diagnostic pathway that may contribute to delays in timely diagnosis. Patients in rural areas with limited access to healthcare resources may present late after symptom onset. There also could be delays in sample dispatch and transportation to national and regional reference laboratories. The reasons for delays may be country and region-specific, and there have been limited studies of YF diagnostic networks in African countries. One analysis found that the lag between symptom onset and specimen collection contributed most significantly to delays in YF diagnosis in the Central African Republic [17]. In contrast, another study found that the lag between specimen collection to shipment to the national reference laboratory was the greatest contributor to delays in Uganda [18]. Thus, there is a need for further studies on the timeliness of YF testing in African countries.

The WHO’s Eliminate Yellow Fever Epidemics (EYE) strategy was launched in 2017 in response to a YF epidemic in Africa that exhausted the global supply of YF vaccines [19]. A key part of the EYE strategy is accurate and timely laboratory diagnosis, following the WHO YF testing algorithm [14,20]. A recent review of laboratory capacity for YF in 25 African countries found that lack of testing resources and shipping capacity limited confirmation of YF cases [12]. The WHO EYE strategy recommends that blood specimens are ideally collected within 14 days of symptom onset, so that RT-PCR can be performed, leading to a faster confirmatory result [14]. In YF high-risk countries, the WHO also recommends specimens to be sent to the national reference laboratory within 3 days of collection for initial testing [14]. While the WHO has established an international YF specimen transport system for shipment to regional reference laboratories with funding from GAVI [21], countries must rely on their own transportation networks for national testing.

To better understand how different steps in the YF diagnostic pathway contribute to the timeliness of YF testing in Ghana, we evaluated the timing of symptom onset, specimen collection, and specimen arrival for testing for suspected YF cases in Ghana during 2018-2022 in relation to WHO metrics. Because of the time it takes for confirmatory testing at the regional reference laboratory, when there are multiple positive YF IgM results at the NPHRL in Ghana, an outbreak response is triggered. Additionally, the NPHRL in Ghana has recently gained the ability for YF RT-PCR testing. Therefore, our analysis focused on the time between symptom onset and specimen arrival for initial YF testing, recognizing its critical role for timely case detection and outbreak response in Ghana.

## Methods

### Setting

The Republic of Ghana is a country in West Africa and part of the WHO Africa Region (AFRO) YF laboratory network. Since 2019, there have been 260 districts and 16 regions in Ghana. The districts and regions in Ghana for this study are shown in S1 Fig. One additional district (Guan) was added in October 2021 and is not included in this analysis.

The NPHRL in Korle Bu has oversight over the zonal public health laboratories and does all YF IgM testing, forecasting, and resource procurement in Ghana. If there is a high index of suspicion for other viral hemorrhagic fevers, RT-PCR testing is done at the Noguchi Memorial Institute for Medical Research. Positive YF IgM specimens are transported to the WHO regional reference laboratory at Institut Pasteur de Dakar, Senegal for confirmatory testing. There have been recent efforts for the NPHRL in Ghana to gain WHO accreditation for YF confirmatory testing.

### Data Collection

Data on YF testing in Ghana from 2018-2022 were collected from the Ghana Health Service’s Epi Info database. We collected data on the reporting health facility, patient’s town of residence and demographics, YF vaccination status, date of symptom onset, date patient was evaluated, date of specimen collection, date of specimen dispatch, date of specimen arrival at the NPHRL, and NPHRL test result. The exact dates of when the tests resulted were unavailable; per national protocols all YF testing at the NPHRL should be done within 72 hours of specimen arrival.

### Statistical Analysis

Our primary outcomes of interest were overall days from symptom onset to specimen arrival at the NPHRL as well as (i) days from symptom onset to specimen collection and (ii) days from specimen collection to arrival at the NPHRL. Records with specimen collection and/or arrival dates occurring a year or more after symptom onset were excluded from our analysis due to suspected data entry errors (see S1 Appendix for cohort selection). We calculated national and district-level descriptive statistics for each of our outcomes. We compared these outcomes to two WHO metrics: the percent of specimens collected within 14 days of symptom onset and the percent of specimens that arrived at the NPHRL within 3 days of collection [14]. We also calculated these metrics pre and post COVID-19 (specimen collection during 2018-2019 and 2020-2022), given the influence the pandemic may have had on testing. Aggregate statistics by district are available online [22]. All statistical analyses and figures were created using R version 4.4.0 [23].

We calculated Moran’s I to determine the degree of spatial autocorrelation among district-level YF testing statistics. District boundaries were obtained from the Ghana Statistical Service (GSS)/Humanitarian Data Exchange (https://data.humdata.org/dataset/cod-ab-gha). Moran’s I was calculated in R using the spdep package, and maps were created using the R packages: sf, ggplot2, viridis, and patchwork. A spatial weights matrix was constructed using queen contiguity and row-standardized weights were used with the assumption that spatial autocorrelation would be most influenced by immediate neighboring districts.

## Results

From 2018-2022, there were 6609 specimens collected from 6447 patients to be tested for yellow fever, and 158 patients had multiple samples collected. The characteristics of the patients tested for YF are shown in Table 1. The majority of patients tested for YF were male (60%), lived in rural areas (74%), and were under 15 years of age (59%). A record of prior YF vaccination was available for 28% of patients. The median age was nine years (range 0-95 years). 74/5142 (1%) of samples were IgM positive for YF.

**Table 1.** Characteristics of patients tested for yellow fever in Ghana (2018-2022)

|  |  | **Total**<br>**(N=6447)** |
| --- | --- | --- |
| **Age (years)** |  |  |
|  | 0-14 | 3809 (59.1%) |
|  | 15-29 | 1406 (21.8%) |
|  | 30-44 | 696 (10.8%) |
|  | 45-59 | 227 (3.5%) |
|  | 60+ | 64 (1.0%) |
|  | Unknown | 245 (3.8%) |
| **Sex** |  |  |
|  | F | 2597 (40.3%) |
|  | M | 3833 (59.5%) |
|  | Unknown | 17 (0.3%) |
| **Location of residence** |  |  |
|  | Rural | 4793 (74.3%) |
|  | Urban | 1333 (20.7%) |
|  | Unknown | 321 (5.0%) |
| **Record of prior YF vaccination** |  |  |
|  | Yes | 1771 (27.5%) |
|  | No | 4676 (72.5%) |

Overall, the median time from symptom onset to specimen arrival at the NPHRL was 10 days (IQR 6-17, range 0-353 days), with 5892/6345 (93%) of specimens meeting the WHO-recommended threshold of collection within 14 days of symptom onset. The median time from symptom onset to specimen collection was 4 days (IQR 2-7 days, range 0-349 days), and from specimen collection to arrival at the NPHRL was 5 days (IQR 2-9 days, range 0-229 days). 2653/6471 (41%) of specimens arrived at the NPHRL within the WHO-recommended threshold of 3 days. Maps depicting the timing of YF specimen collection and arrival for testing shown in Fig 1 and Fig 2.

**Fig 1.**
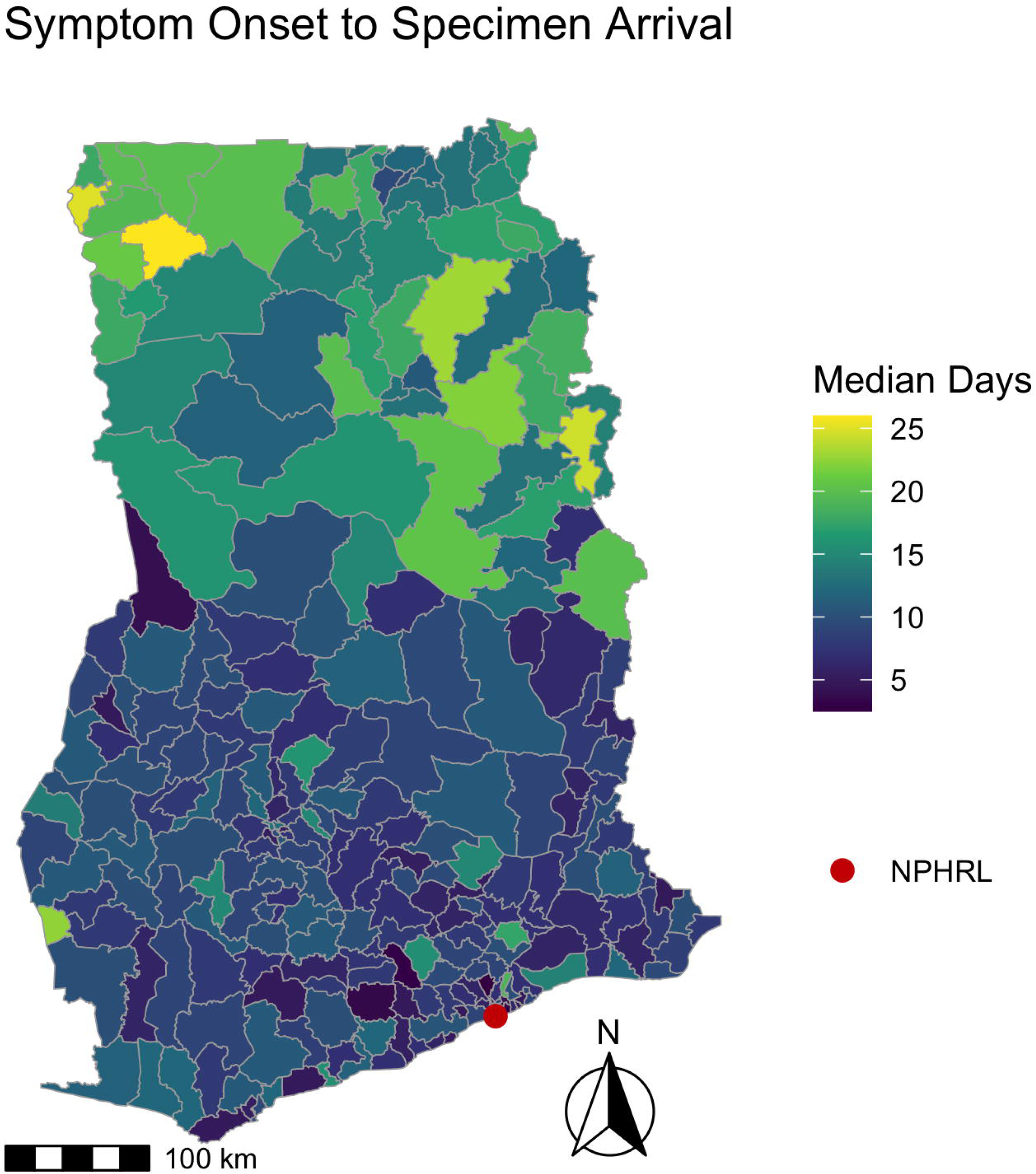
Time between symptom onset to specimen arrival for yellow fever testing by district in Ghana 2018-2022. For suspected YF cases from 2018-2022, the map depicts the median days for each district from symptom onset to specimen arrival at the NPHRL. The shapefile layers used to create the map were obtained from the Ghana Statistical Service (GSS)/Humanitarian Data Exchange (https://data.humdata.org/dataset/cod-ab-gha).

**Fig 2.**
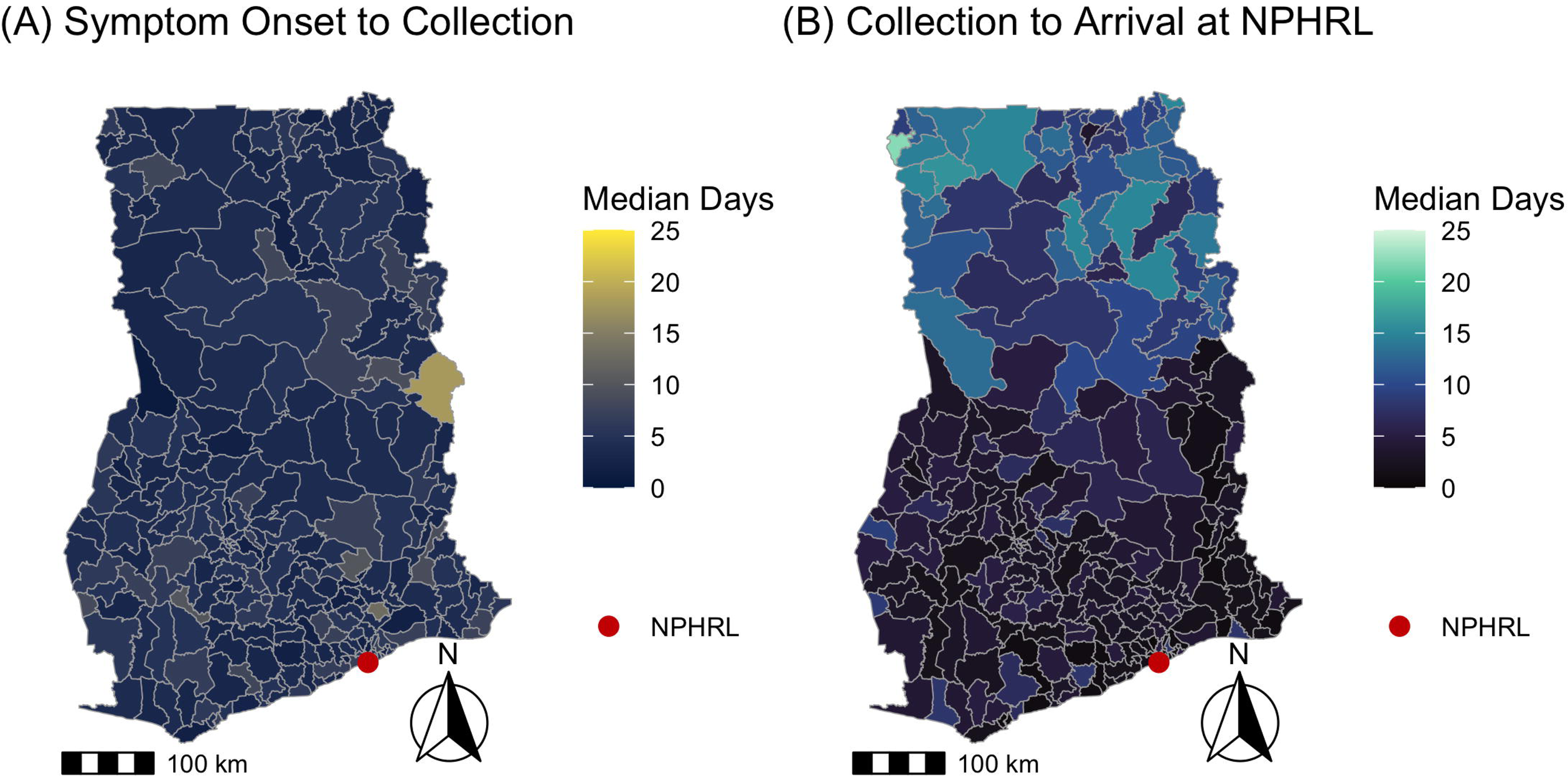
Timing of yellow fever specimen collection vs specimen arrival by district in Ghana 2018-2022. For suspected YF cases from 2018-2022, the maps depict the (A) median days from symptom onset to specimen collection vs (B) median days from specimen collection to specimen arrival at NPHRL. The shapefile layers used to create the maps were obtained from the Ghana Statistical Service (GSS)/Humanitarian Data Exchange (https://data.humdata.org/dataset/cod-ab-gha).

The full descriptive statistics comparing the entire study time period (2018-2022) to before and after COVID-19 (2018-2019 vs 2020-2022) are in the S1 Appendix. From 2018-2019, 1936/2087 (93%) of specimens were collected within 14 days of symptom onset, and 1097/2107 (52%) arrived within 3 days of collection. During 2020-2022, 3956/4258 (93%) of specimens were collected within 14 days of symptom onset, and 1556/4364 (36%) arrived within 3 days of collection. A higher proportion of specimens arrived within 3 days of collection during 2018-2019 compared to 2020-2022 (two-sample Z test of proportions, p < 0.0001).

The West Gonja district, where the 2021-2022 YF outbreak originated, had the highest number of patients tested for YF (161). Fig 3 shows the number of patients tested for YF per district in 2018-2022.

**Fig 3.**
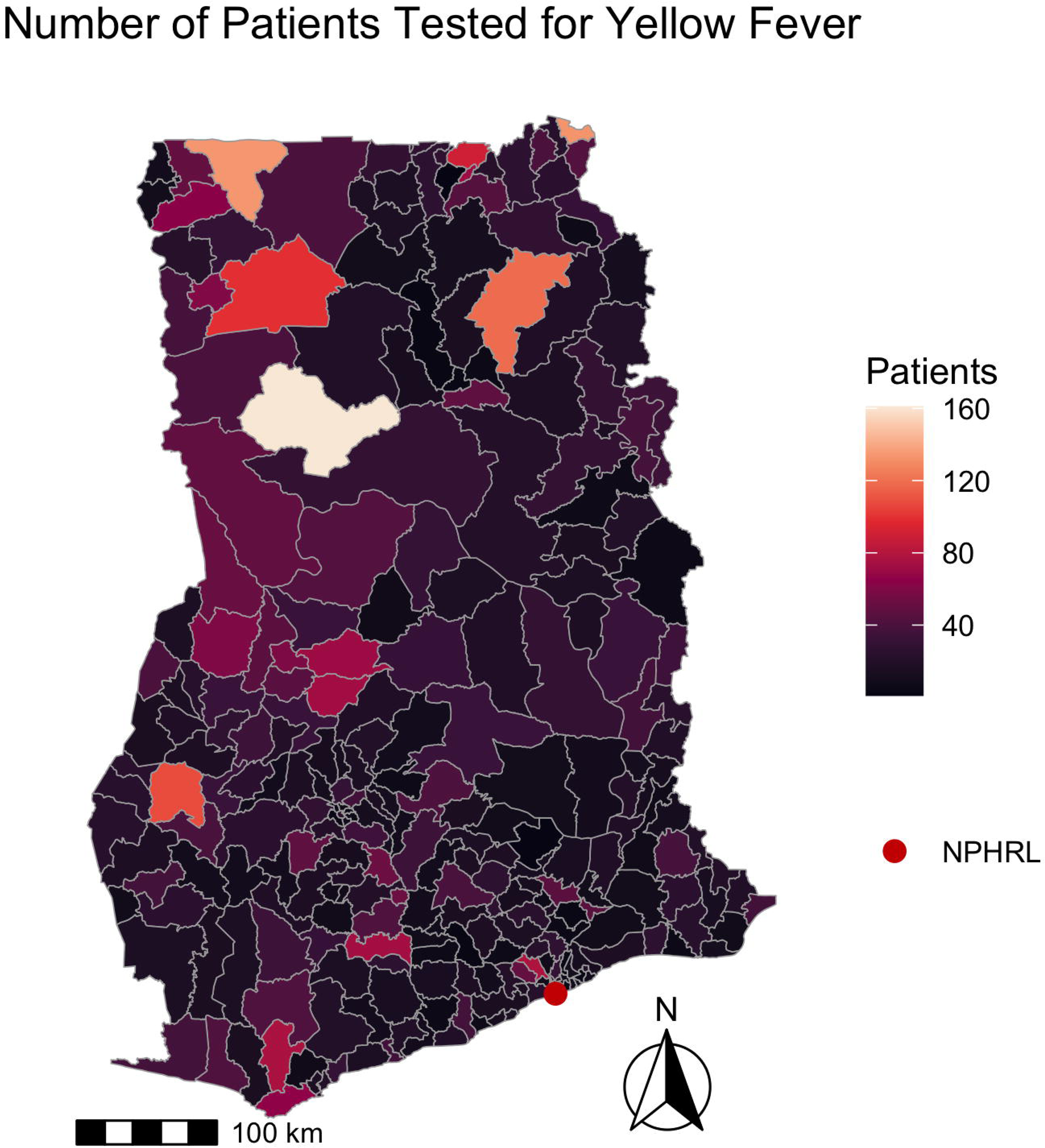
Number of patients tested for yellow fever by district in Ghana 2018-2022. The map depicts the number of patients tested for yellow fever by district from 2018-2022. The shapefile layers used to create the map were obtained from the Ghana Statistical Service (GSS)/Humanitarian Data Exchange (https://data.humdata.org/dataset/cod-ab-gha).

There was significant spatial autocorrelation among districts regarding the number of patients tested (Moran’s I = 0.15, p <0.001), the median days from symptom onset to specimen arrival (Moran’s I = 0.52, p <0.001), and median days from specimen collection to arrival (Moran’s I = 0.69, p <0.001). There was no significant spatial autocorrelation among districts regarding the median days from symptom onset to specimen collection (Moran’s I = 0.03, p = 0.22).

## Discussion

We found that the timing of specimen collection and arrival for YF testing varies across districts in Ghana. Nearly all samples (93%) in our study were collected within the WHO-recommended time frame of 14 days from symptom onset, during which RT-PCR can theoretically be performed. In contrast, we observed delays between specimen collection and arrival at the NPHRL for testing, with only 41% of specimens arriving within the WHO-recommended three days. These findings suggest targets for improving YF diagnosis at a country level in Ghana, specifically highlighting the importance of timely specimen transport.

Our observed duration (median four days) from symptom onset to specimen collection was similar to that seen in Uganda from 2017-2022 (median three days) [18]. In contrast, the time from symptom onset to specimen collection for YF contributed to 60% of the lag time in the Central African Republic from 2007-2012 [17] and was also higher during the 2016 YF outbreak in the Democratic Republic of the Congo (median 9 days) [2].

Our finding of frequent delays from specimen collection to arrival at NPHRL was also consistent with findings in Uganda [18], and likely reflects the structures of YF diagnostic networks in these countries. For example, in Ghana specimens from suspected YF cases are often batched prior to transportation to the NPHRL. This batching process likely contributes to the lag in specimen arrival for testing. We also found that timely arrival of specimens to the NPHRL decreased by roughly a third during 2020-2022, which could reflect the impact of COVID-19 in straining the YF diagnostic network.

We also identified district-level patterns in the timeliness of YF testing in Ghana. Neighboring districts were more likely to have similar YF testing volume and time from specimen collection to arrival at the NPHRL. In all of the districts in the northernmost regions in Ghana (Upper West, Upper East, Savannah, Northern, and Northern East), the median time from specimen collection to arrival at the NPHRL was delayed beyond the WHO’s recommended threshold of within 3 days. This suggests that improving YF specimen transport or establishing testing closer to districts that are far from the NPHRL could improve timeliness of YF diagnosis and outbreak detection.

Given multiple recent outbreaks of viral hemorrhagic fevers, including YF, in Africa, there has been a recent emphasis on improving early detection and outbreak response [24–26]. Organizations such as the WHO and FIND have assessed accessibility and optimizing diagnostic networks for infectious diseases [27]. The 7-1-7 strategy used by Resolve to Save Lives emphasizes outbreak detection within 7 days, public health notification within 1 day, and an initial response within 7 days [28]. These metrics as well as the WHO’s performance indicators can help guide timely YF diagnosis and response.

A limitation to our analysis is that we were unable to confirm the timing of when YF tests resulted or were reported to district health facilities. From interviews with NPHRL staff, we learned that there may be an inadequate supply of reagents for local YF IgM MAC-ELISA testing, and testing support from the WHO primarily occurs during outbreaks. Therefore, it is possible that resource constraints at the NPHRL may also contribute to delays in testing. We also learned that the confirmatory testing at the regional reference laboratory in Senegal could contribute further to diagnostic delays given the additional costs for transportation and prolonged turn-around time. However, given that local public health responses for YF in Ghana have been initiated with clusters of YF IgM positivity and there is new capacity for YF RT-PCR at the NPHRL, the timing of initial testing in Ghana will be most important for local outbreak detection.

Our results suggest important targets for strengthening the YF diagnostic cascade in Ghana. Specifically, facilitating faster specimen transport could be prioritized. Ghana has national diagnostic transportation systems for HIV and tuberculosis, and such a system for YF could address delays inherent in batching samples. A pilot study in Mali showed that using a trained postal service improved timeliness for YF specimen arrival, however it also increased costs 8-fold [29]. Therefore, costs for a national YF transportation system in Ghana may be prohibitive, so it is important to consider alternative diagnostic strategies such as decentralized testing and point-of-care testing.

Considering decentralized testing, the Zonal Public Health Laboratory in Tamale is closer to the northern districts and could be an ideal location for expanded YF testing given recent outbreaks in this area [5]. Alternatively, point-of-care tests, such as a lateral flow YF IgM assay that was recently validated in Ghana [30], could be used in remote districts. Such strategies could be especially important for timely case detection during outbreaks. For example, a mobile lab was deployed for YF testing during the 2016 YF epidemic in the Democratic Republic of the Congo with positive results [31]. While our study is specific to Ghana, our findings could be applicable to many other countries in Africa that have centralized testing for YF near urban capitals but have outbreaks originating in remote rural areas.

Analyzing national level diagnostic networks for YF will be essential for preparing and responding to future YF outbreaks. The timing of YF specimen arrival at the NPHRL in Ghana contributes to delays in testing, especially among more remote northern districts that could be at greater risk for YF outbreaks. Strengthening specimen transport systems and/or decentralized testing could improve timely detection of YF, leading to quicker patient isolation, supportive care, vector control, and vaccination campaigns.

## Data Availability

Aggregate data are available at: doi:10.6084/m9.figshare.29265026.v1

https://figshare.com/articles/dataset/_b_Timeliness_of_Yellow_Fever_Specimen_Collection_and_Transport_in_Ghana_2018-2022_Dataset_b_/29265026

## Ethical approval

This study was approved as part of the Ghana Laboratory Network Project by the Ghana Health Service Ethical Review Committee.

## Acknowledgements

The authors would like to thank their colleagues from the Ghana Laboratory Network Project, Ghana Health Service, University of Ghana, and Noguchi Memorial Institute for Medical Research.

## Author contributions

Conceptualization, Analysis, Writing-original Draft: SDJ

Data curation: HG, EK, RO, JFD

Writing-reviewing and editing: SDJ, DWD, LFS, FA, EK, DL, GB, HG, RO

Funding: SDJ, DWD, LFS, EK

## Supporting Information

## S1 Appendix

**S1 Fig. Ghana Districts and Regions 2018-2022**

The 260 districts and 16 regions in Ghana that were analyzed for yellow fever testing from 2018-2022 are shown with the National Public Health and Reference Laboratory (NPHRL).

