## Supplementary material for "Timeliness of Yellow Fever Specimen Collection and Transport in Ghana, 2018-2022": S1 Appendix

**Cohort Selection of Specimens for Analysis of Timeliness**

**Sensitivity Analysis**

Our primary analysis included suspected YF cases from 2018-2022. We also conducted a sensitivity analysis comparing testing before and after the COVID-19 pandemic (2018-2019 vs 2020-2022) given the potential impacts on testing.

|  | Overall 2018-2022 | 2018-2019 | 2020-2022 |
| --- | --- | --- | --- |
| Days from Symptom Onset to Specimen Received | Median 10 (IQR 6-7, Range 0-353) | Median 9 (IQR 6-14, Range 0-296) | Median 11 (IQR 7-18, Range 0-353) |
| Days from Specimen Collection to Received | Median 5 (IQR 2-9, Range 0-229) | Median 3 (IQR 2-7, Range 0-94) | Median 6 (IQR 2-10, Range 0-229) |
| Days from Symptom Onset to Specimen Collection | Median 4 (IQR 2-7, Range 0-349) | Median 4 (IQR 2-7, Range 0-276) | Median 4 (IQR 2-7, Range 0-349 |
| <14 days to Specimen Collection | 5892/6345 (93%) | 1936/2087 (93%) | 3956/4258 (93%) |
| <3 days to Specimen Arrival | 2653/6471 (41%) | 1097/2107 (52%) | 1556/4364 (36%) |
