## Supplementary material for "Timeliness of Yellow Fever Specimen Collection and Transport in Ghana, 2018-2022": S1 Fig

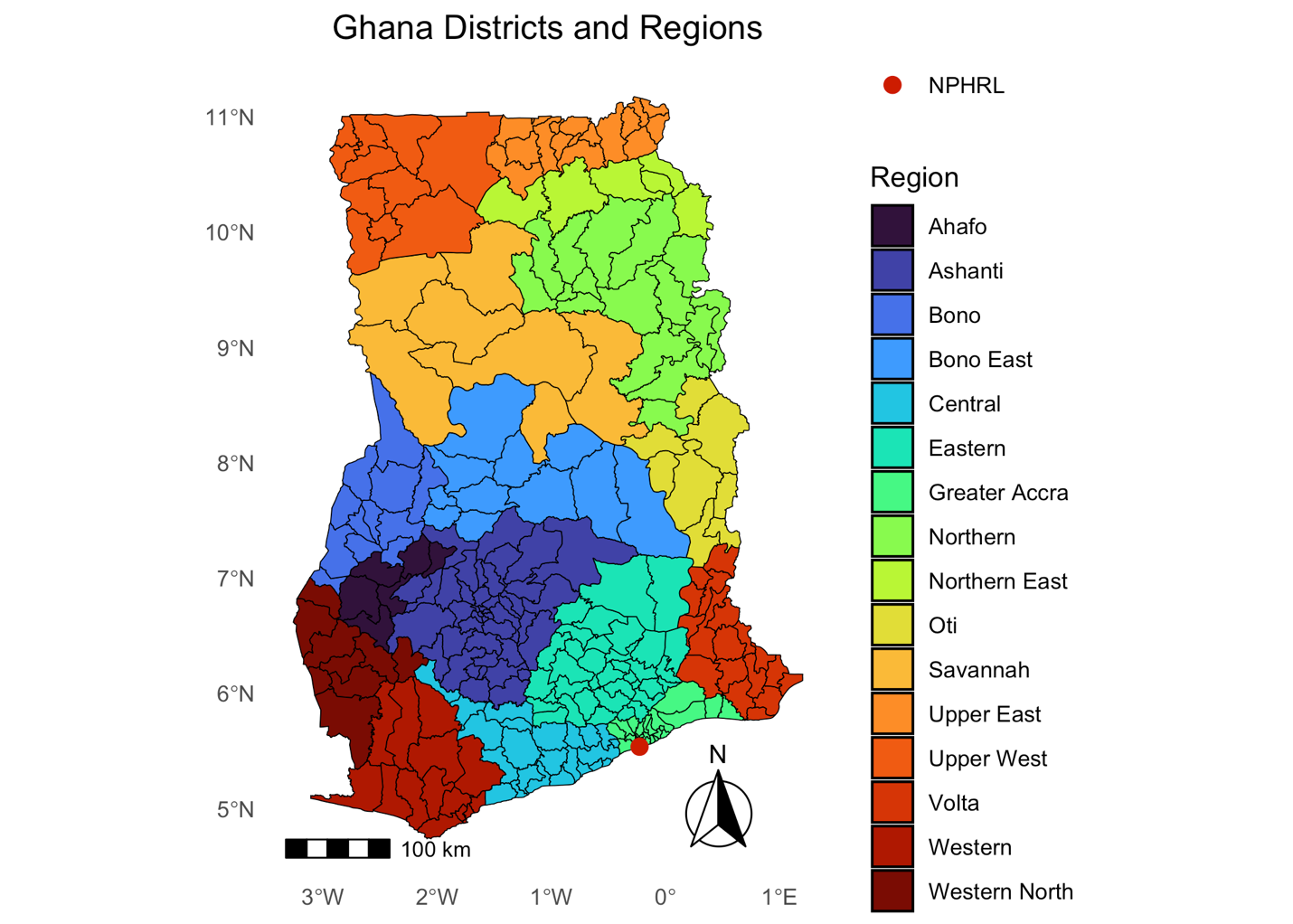

**S1 Fig. Ghana Districts and Regions 2018-2022**

The 260 districts and 16 regions in Ghana that were analyzed for yellow fever testing from 2018-2022 are shown with the National Public Health and Reference Laboratory (NPHRL).
